# Labored breathing pattern: an unmeasured dimension of respiratory pathophysiology

**DOI:** 10.1101/2024.01.27.24301872

**Authors:** Valerie E. Cyphers, Swet M. Patel, Brendan D. McNamara, William B Ashe, Sarah J. Ratcliffe, J. Randall Moorman, Jessica Keim-Malpass, Shrirang M. Gadrey, Sherry L. Kausch

## Abstract

**Introduction:** Respiratory failure is a common organ failure syndrome in hospitalized patients^1^. Vital sign monitoring (like respiratory rate & oximetry) is a necessary aspect of risk stratification, but it is not sufficient. In one study of hospitalized patients, 46% of the patients had no significant vital sign change in the 24 hours before an unplanned intubation^2^. Therefore, clinicians must also monitor for physical diagnostic signs that link the appearance of breaths to respiratory instability. Many pathognomonic patterns of high-risk labored breathing have been described. For example, when rib-dominant breaths alternate with abdomen-dominant ones, the patient is said to exhibit “respiratory alternans”, a sign of inspiratory muscle overload^3^. However, the manual assessment of such signs lacks sensitivity, inter-rater reliability, and scalability^4^. We sought to (a) identify technologies that can measure labored breathing and (b) assess their readiness for clinical adoption by hospitals.

**Methods:** We selected four well-established diagnostic signs of labored breathing: (1) respiratory rate variability, (2) recruitment of accessory muscles (upper-rib elevation by the scalene and sternocleidomastoid muscles), (3) Abdominal Paradox (rib-abdomen asynchrony), and (4) respiratory alternans (rib-dominant breaths alternate with abdomen-dominant ones). We systematically searched PubMed using pre-specified keywords corresponding to these four signs. We identified 2868 abstracts. Two reviewers independently screened each abstract to ensure that it reported on technology that quantified the diagnostic sign of interest. A third reviewer resolved any disagreements. We excluded 2423 articles with an abstract review and included 445 articles for full paper review. We excluded an additional 127 articles after full paper review, and we were unable to acquire 4 articles. We included the remaining 314 articles for analysis.

**Results:** Quantification of labored breathing has been attempted for over 50 years; the earliest study included in our analysis was published in 1975. Over 30 different hardware configurations have been tried, either alone or in combination; but none of them has been validated as a comprehensive solution to measure all the four diagnostic signs that we studied. Despite enormous improvements in sensor technologies and computing capacity, the scale of investigation has not meaningfully increased since 1975. In the first decade of kinematic measurements (1975–1984), there average annual number of studies was 2.7 and the median sample size was 19. In the decade prior to our study (2013–2022), the average annual number of studies was 11.3 and the median sample size was 20. To this day, a majority of the studies are conducted in a specialized laboratories (73% between 2013-2022) rather than clinical practice settings. Most studies aimed to measure the construct validity of a technology (19%) or to describe kinematic distributions in specific clinical scenarios (77%). Rarely did studies attempt to quantify the predictive validity for a clinical outcome (4%). We did not find any clinical trial where a kinematics-based early warning intervention was tested.

**Conclusions:** This study describes a major bottleneck in the translation of bedside diagnostic signs of high-risk labored breathing patterns into measurable physiomarkers of respiratory instability. Despite half a century of attempted measurement, the technology readiness level for clinical adoption remains low.

## Introduction

Respiratory failure is the most common organ failure syndrome in US hospitals^1^. It necessitates endotracheal intubation for invasive mechanical ventilation in about 800,000 hospitalizations annually^5^. It’s estimated national costs were $27 billion per year, representing 12% of all hospital costs^5^. The COVID-19 pandemic further increased the incidence of severe respiratory failure and the nationwide demand for ventilators^6^. Respiratory failure requiring mechanical ventilation is associated with an in-hospital mortality rate of 35%^5^. Among emergency intubations for unanticipated deterioration, the mortality rate can approach 50%^2^. Up to 40% of all in-hospital cardiac arrests are linked to respiratory deterioration^7^.

One half of all in-hospital intubations are performed during the initial presentation to the emergency room^8^. The other half are performed during the hospitalization. In these cases, the patient suffers a clinical deterioration while in the hospital which precipitates intubation. For early detection of respiratory deterioration, hospitalized patients are monitored by many physiological “track and trigger” warning systems, but the sensitivity of such systems is poor^9^. Too often, subtle pathophysiologic antecedents of instability fail to trigger early alerts^10^. Delayed calls to emergency response teams are common, and independently predict higher mortality^11^. Abrupt respiratory compromise causes 64,000 unplanned intubation events every year in US hospitals^2^. Predictive analytics that use conventional physiomarkers (respiratory rate, oximetry, heart rate, blood pressure etc.) have not revealed a reliable signature of respiratory instability. In one review of 448 unplanned intubation events, 46% of the events were not preceded by any significant change in any vital sign^2^. Even when vital signs did change significantly, their trajectory was complex and heterogeneous. For example, pulse oximetry saturation dropped significantly in 7% of patients, but rose significantly in 6%^2^.

To improve advance warning systems for respiratory deterioration, researchers are applying increasingly sophisticated modelling techniques (regression, decision trees, clustering, and deep learning) to increasingly complex datasets^12–14^. Modern datasets include the basic vital signs like respiratory rate and oximetry, basic electronic health record (EHR) variables like laboratory tests and diagnosis codes, continuous monitoring signals like electrocardiography and impedance pneumography^15,16^, and advanced bioinformatic variables (like natural language processing of notes^17,18^. However, the real-time implementation of advanced (often proprietary) predictive models is significantly more costly than that of a simple, open-source early warning score (like NEWS). The costs include data storage, computation capacity, licensing fees, and human responders to alarms. If the clinical gains are incremental, or if they are offset by a manifold rise in false alarms, then cost-benefit metrics (e.g., number needed to monitor) do not justify the upgrade. The economic barriers to adoption are particularly strong in underprivileged health systems and the benefits of complex analytics are often inaccessible. In most settings, therefore, respiratory remains difficult to predict.

One way in which clinicians manage this risk is to visually inspect their patients’ breathing motion patterns. This idea is rooted in the fundamental principles of bedside diagnosis that link the appearance of breaths (i.e., respiratory kinematics) to respiratory pathology^19^. For example, when rib-dominant breaths alternate with abdomen-dominant ones, the patient is said to exhibit “respiratory alternans” which is a manifestation of inspiratory muscle overload^3^. Many such pathognomonic visual signs are known to clinicians, but there has been no way to quantify them in routine practice. Clinicians use qualitative visual inspections to assess for abnormal breathing patterns. Such assessments lack sensitivity and inter-rater reliability^4^, and are manually effort-intensive. Therefore, we sought to (a) identify technologies that can measure labored breathing and (b) assess their readiness for clinical adoption by hospitals.

## Methods

We reviewed textbooks of physical diagnosis^20^, pertinent expert commentaries^3^, research studies, and used our own clinical domain expertise to identify the breathing motion patterns that are most widely used as indicators of respiratory instability. We excluded respiratory rate because it is a conventional vital sign and it’s measurement is a well-addressed in the literature^21^. We identified four labored breathing patterns that adequately represent the type of measurements of interest to clinicians: (1) irregular respiratory rhythm or elevated respiratory rate variability where one sees short breath intervals alternate with long ones, (2) recruitment of accessory muscles of respiration where one can observe increased contraction of neck muscles (like sternocleidomastoid and scalene) which elevate the upper ribs, (3) asynchronous abdominal motion, whereby synchrony between the chest and abdomen is lost (in the most severe form, known as abdominal paradox), and (4) respiratory alternans, during which breathing switches between being chest-predominant and abdomen-predominant phenotypes.

We systematically searched PubMed using pre-specified keywords corresponding to these four breathing patterns. Al articles published before our search (June 2022) were included. Specific search terms are outlined in Table 1. We used the following inclusion criteria: (a) study describes a technology to measure one or more of the specified breathing patterns in human subjects, (b) the described technology is applicable in spontaneously breathing individuals, (c) study reports measurements made during an unmodified respiratory cycle. We excluded studies where a ventilator or other respiratory support device was used to collect respiratory data and/or subjects were only assessed during modified breathing patterns such that tidal breaths were not captured (like coughing, singing, or forced expiration with breath hold).

We used the systematic review program Rayyan for screening and decision tracking. In a blinded screening process, two reviewers (VEC, SMP, BDM) independently assessed each abstract to identify studies that met the pre-specified eligibility criteria. A third reviewer (SMG) resolved any disagreements. Reviewers cross-screened for the other respiratory signs of interest when reviewing abstracts for any given keyword search. For example, if the search for respiratory rate variability returned an abstract that met eligibility for respiratory alternans, then this was included in final analysis. Following the abstract screening phase, one reviewer (VEC) performed full text analysis of the papers that met inclusion criteria to confirm eligibility and extract data. The datapoints recorded during full-text review were (a) year of publication, (b) study size (number of subjects, not number of measurements), (c) study setting (a clinical practice setting versus specialized laboratory; type of practice setting or laboratory); (d) stage of technology validation (construct validity, descriptive study, predictive validity, clinical trial).

## Results

The results of the outlined selection process are shown in Figure 1. The electronic search resulted in 2,868 abstracts. Based on title and abstract, 2,407 articles were eliminated because they did not meet eligibility criteria. The remaining 461 abstracts corresponded to 445 separate articles. We excluded an additional 127 articles after a review of the full article, and we were unable to acquire 4 articles. We included the remaining 314 articles for data abstraction (a full list of studies is presented in the supplemental file).

**Figure 1:**
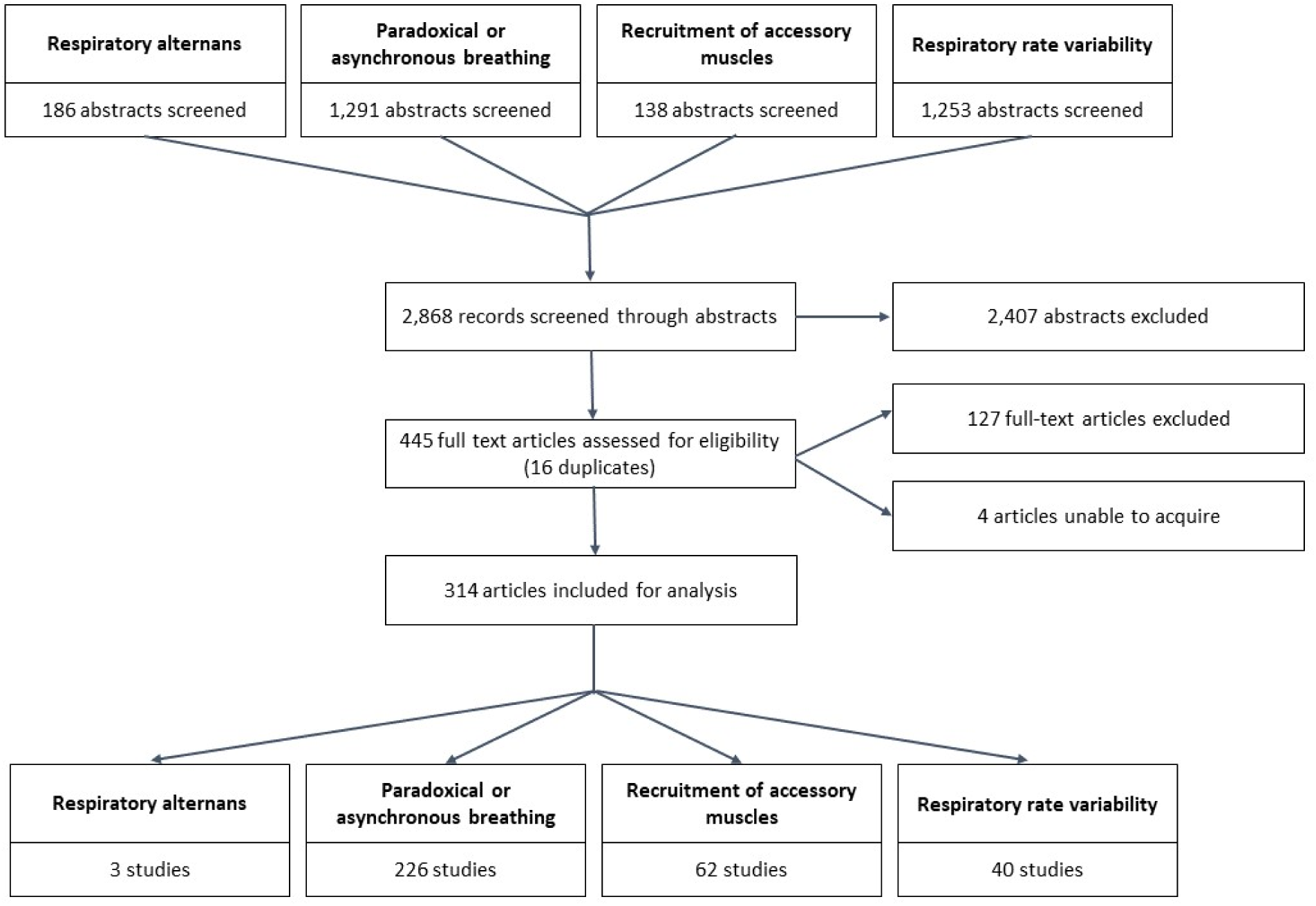
Screening and full article reviews. This figure shows the number of studies that were included in the abstract screening (2868), full article review (445), and data collection (314) stages of our study. We note that the study numbers in the bottom four boxes adds up to 331 instead of 314 because 17 studies measured 2 signs of labored breathing and were counted in two boxes.

The earliest study included in our analysis was published in 1975. We identified over 30 types of devices that have been applied for the quantification of labored breathing, either alone or in combination. We categorized them, as shown in Figure 2, based on the sensor type. The most frequently used sensors measured inductance / strain (e.g., respiratory inductance plethysmography), followed optical (e.g., optoelectronic plethysmography), electromyography, and inertial (e.g., accelerometer) sensors. No system has been validated as a measurement for all four signs of labored breathing that we studied.

**Figure 2:**
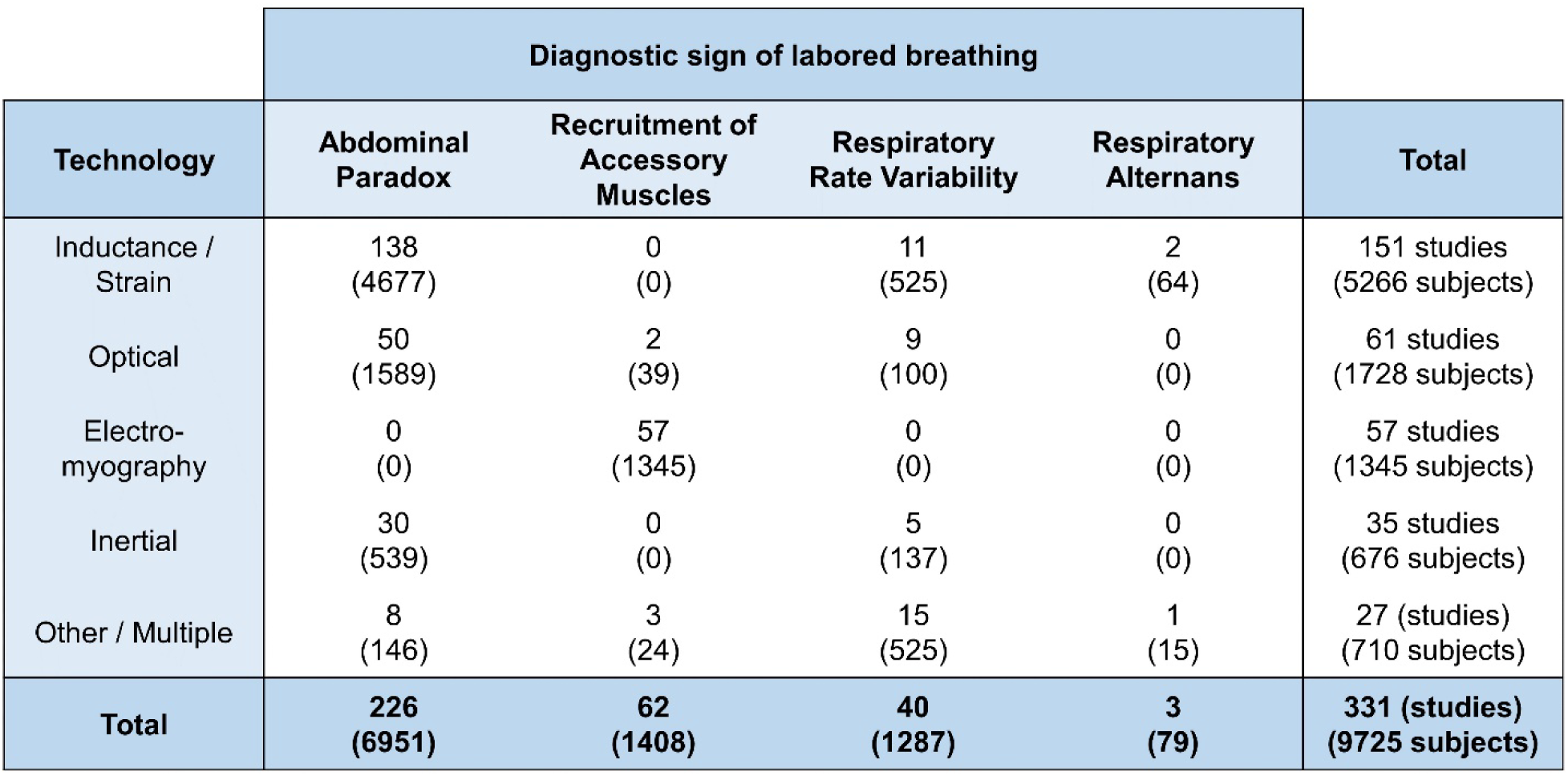
Technologies for labored breathing measurement Some sensor-metric combinations were seen frequently. For example, inductance plethysmography for the measurement of abdominal paradox (rib-abdomen asynchrony) is the single most frequent type of labored breathing measurement. However, no technology had been validated as a method for comprehensive assessment of labored breathing. (Of note, 17 studies measured two diagnostic signs. These were counted twice in this tabulation, making the total 331 instead of 314.)

Despite major advances in sensors and computers between 1975 and 2022, the scale of studies quantifying breathing patterns had not increased (Figure 3). We identified only 314 studies that had ever quantified the signs of labored breathing that we studied (6-7/year). To this day, most studies are conducted in specialized laboratories.

**Figure 3:**
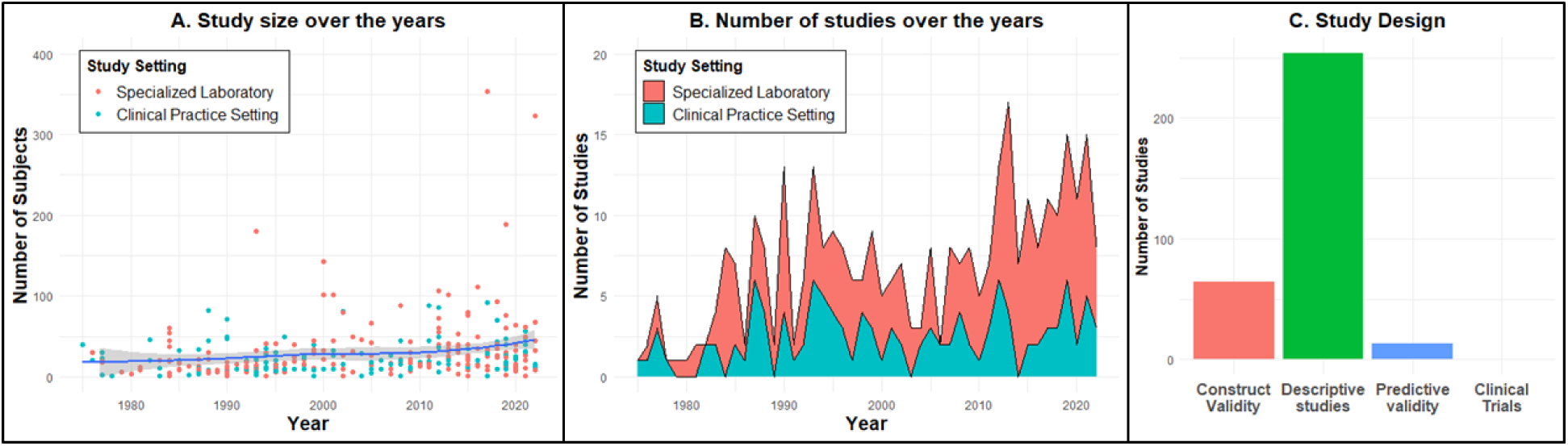
Technology readiness for clinical adoption remains low. This figure depicts the magnitude and type of research that has focused on quantification of labored breathing patterns. Panels A & B show that the scale of investigation has not meaningfully increased since 1975. In the first decade of kinematic measurements (1975–1984), there average annual number of studies was 2.7 and the median sample size was 19. In the decade prior to our study (2013–2022), the average annual number of studies was 11.3 and the median sample size was 20. To this day, a majority of the studies are conducted in a specialized laboratory (73% between 2013-2022). Panel C shows a bottleneck in the bench to bedside spectrum. Most studies aim to measure the construct validity of a technology (19%) or to describe kinematic distributions in specific clinical scenarios (77%). Rarely do studies attempt to quantify the predictive validity for a clinical outcome (4%). We did not find any clinical trial where a kinematics-based early warning intervention was tested. Therefore, we conclude that the technology readiness level for clinical adoption remains low.

## Discussion

We conducted a scoping review of technologies that can quantify four major breathing motion abnormalities. We found a critical bottleneck in the translation of bedside diagnostic signs of labored breathing into measurable physiomarkers of respiratory instability. Despite multiple potential technologies being tried since 1975, their readiness for clinical adoption remains low.

Physician ratings of labored breathing patterns have been shown to improve predictions of intubation beyond the conventional predictors^22^. In the absence of quantitative measurements, such insights are confined to conventional clinical wisdom; they have not been integrated into predictive analytics. For an informative clinical sign to be integrated into predictive analytics, it has to be measurable with ease, accuracy, and reproducibility. For example, the respiratory rate was included in the NEWS (National Early Warning Score) model after an analysis of 200,000 respiratory rate values across 35,585 hospitalizations^23^. Similarly, it was included in the qSOFA (quick Sequential Organ Failure Assessment) model after an analysis of respiratory rates across 2 million encounters in 175 world-wide hospitals^24^. Our study revealed that labored breathing patterns had never been quantified on a scale needed for predictive modelling. Most prevailing ideas about the significance of breathing patterns are based on experts’ opinions or small studies in research settings. Little is known about the real-world incidence of any breathing motion abnormality, or its sensitivity, specificity, or predictive value for respiratory failure.

Crucially, breathing pattern abnormalities report a unique dimension of physiology that is not captured by the conventional vital signs. In a prospective study of 1,134 subjects, only a small portion of the variance in physicians’ rating of labored breathing was explained by vital signs and laboratory data^22^ This is important because the performance of a predictive algorithm or early warning score is fundamentally linked to the quality of data on which it is trained and applied. If the conventional data sources fail to capture a certain portion of pathophysiologic information, then that missing information restricts the performance of any model trained on the data. As such, simpler models based on complete physiological information might to outperform complex models based on partial information. Improvements in technologies that quantify labored breathing can greatly improve the early detection of respiratory deterioration. Therefore, a solution for the problem identified by our study is urgently needed.

## Data Availability

All data produced in the present study are available upon reasonable request to the authors

