## Supplemental file for "Labored breathing pattern: an unmeasured dimension of respiratory pathophysiology"

**Supplemental Content**:

Table 1: PubMed Search Details

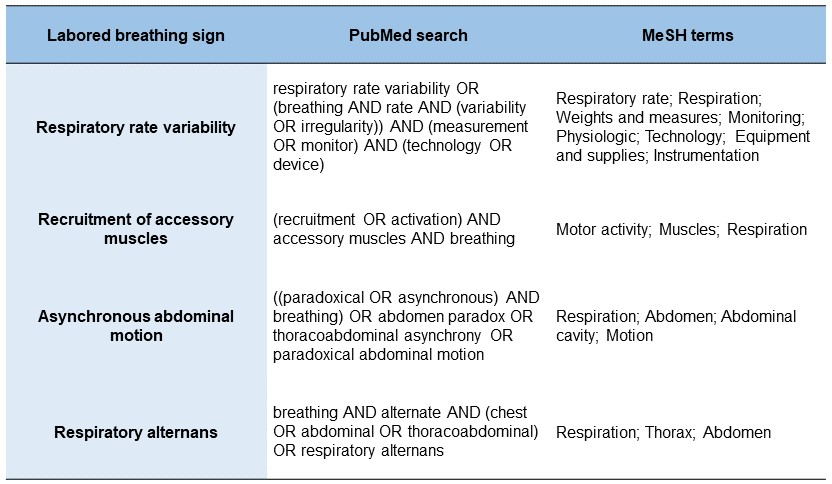

**Studies included in analysis**
